# Rapidly deployable mobile BSL-3 laboratory: A response to nipah virus outbreak in Kozhikode, Kerala, India 2023

**DOI:** 10.1101/2024.06.06.24308303

**Authors:** Rima R. Sahay, Deepak Y. Patil, Anita M. Shete, Sreelekshmy Mohandas, Nivedita Gupta, Devendra T. Mourya, Pragya D. Yadav

## Abstract

The Nipah virus (NiV) outbreak was declared in Kozhikode district, Kerala state, India on September 12, 2023. The local, state, and national authorities worked in an integrated way to tackle and control the outbreak. Indian Council of Medical Research (ICMR) deployed a team from ICMR-National Institute of Virology (NIV), Pune, India along with an indigenously developed and validated Mobile BSL-3 (MBSL-3) laboratory for providing onsite NiV diagnosis. The Kozhikode district of Kerala state has been an epicenter of three NiV outbreaks in May 2018, August 2021, and recently in September 2023. The Ernakulam district, Kerala also reported NiV outbreak in June 2019. In the 2023 outbreak, six confirmed NiV cases were detected with two deaths. During previous outbreaks in 2019 and 2021, the team from ICMR-NIV, Pune had successfully established the field laboratory utilizing the BSL-2 facility for NiV onsite diagnosis. The BSL-3 personnel protective equipment and standard operative procedures were used to handle the clinical specimens. Post COVID-19 pandemic, under the pioneering initiative of the Government of India, ICMR and Klenzaids Contamination Control Pvt Ltd, Mumbai developed a rapidly deployable, pragmatic, access control and containment laboratory on bus chassis. This MBSL-3 laboratory was utilized for the NiV onsite diagnosis for early containment of outbreak reducing the turnaround time to diagnosis to just 4 hours. MBSL-3 laboratory played a significant role in NiV outbreak response and could be utilized in the future also reaching the remotest areas of the country.

## 1. Introduction

Rapid Response Mobile Laboratories (RRMLs) are an innovative approach and advancement in the field of outbreak investigations.^1^ This ‘laboratory on wheels’ setup is controlled through an intelligent control automation system, which maintains the working environment under negative air pressure, maintains equipment parameters, and records all necessary data. Mobile laboratories provide rapidly deployable, effective field diagnostic capabilities for unusual outbreak investigation and medical surveillance. The deployment of mobile laboratories can leverage the capacity of national public health by bringing laboratory resources to remote and inaccessible areas for the on-site early detection of the agent and quick containment.^2^ These containment laboratories works on the mode of ‘ready to go and do’ with the trained personnel even in the remotest areas, challenging situations, with minimum or no risk to the individual, community or environment. The classification of RRML includes five levels: Type I (very compact), Type II (box-based), Type III (medium-scale), Type IV (large-scale), and Type V (full-scale).^1^ The classification is divided into three layers: establishing the basic criteria and features that would apply to all RRML, defining RRML capacity and efficiency, and ensuring response adaptability, seamless integration, and scalability.

## 2. Methods

Under the pioneering initiative of Pradhan Mantri-Ayushman Bharat Health Infrastructure Mission of Government of India, Indian Council of Medical Research (ICMR), partnering with the industrial company Klenzaids Contamination Controls Private Limited, Mumbai developed the first of its kind, field deployable, indigenous Mobile Biosafety level-3 (MBSL-3) laboratory (Type-IV RRML). The laboratory is built on heavy duty vehicle chassis of Bharat Benz with Bharat Stage Emission Standards 6 (BSVI) and equipped with advanced heating, ventilation and air conditioning (HVAC) system with High efficiency particulate air (HEPA) filters which could be operated with electric power or diesel generator. The biological waste management are through double door autoclave and biological liquid effluent decontamination (BLED) system.^3^ In addition, the MBSL-3 laboratory is well equipped with the walk-through shower, dynamic pass box, rapid transfer port for material transfer, advanced communication system, the access control entries, geo-monitoring in built system, availability of petrol generators, and uninterrupted power supply system. The MBSL-3 laboratory was validated by ICMR-National Institute of Virology (NIV), Pune in 2022 by performing onsite and offsite assessments of laboratory equipment and testing, functioning of major installations and decontamination methods as per the defined standard operating procedures (SOPs).

### 2.1. Local Setting

The Kerala state of India, had confirmed its fourth nipah virus (NiV) outbreaks since 2018. The first outbreak was detected in Kozhikode district in May 2018 with total of 18 NiV confirmed cases with 16 deaths.^4^ Further, the second outbreak was detected in Ernakulam district, Kerala in June 2019 with single case non-fatal case.^5^ The third outbreak was again in Kozhikode district in August 2021 which was restricted to one fatal NiV case.^6^

In case of higher number of clinical specimens for testing during an outbreak/epidemic, the MBSL-3 laboratory could be utilized for sample handling, and aliquoting. Post inactivation of the samples in MBSL-3 laboratory, it can be further tested at BSL-2 level laboratory for fast and prompt diagnosis.

On September 12, 2023, Kozhikode district of Kerala, India reported an outbreak of nipah virus.^7^ The state of Kerala faced an uphill task for swift and effective response for containment of this outbreak. As an agile and proactive approach for this public health crisis, the indigenously developed and validated MBSL-3 laboratory was deployed immediately for the first time for enhancing the diagnostic capabilities at the epicentre of the outbreak on September 13.

### 2.2. Relevant changes

The early containment responses of NiV outbreaks depends primarily on the isolation of the confirmed cases and this can only be achieved by quick diagnosis on the clinical specimens. Immediately, following the NiV outbreaks in 2019 and 2021, the trained team from the Maximum Containment Facility of ICMR-NIV, Pune established the onsite field laboratory utilizing the BSL-2 facility with BSL-3 personal protective equipment (PPEs) and the SOPs of biosafety and testing. At that time, as ICMR did not have MBSL-3 laboratory, the on-site field laboratory at BSL-2 level was operationalized for prompt and early diagnosis.

Many biosafety and biosecurity challenges were faced during the onsite field BSL-2 laboratory setup. Setting up a safe laboratory in BSL-2 facility with BSL-3 PPEs for aliquoting nipah suspected samples is a complex task during an outbreak. Outbreaks often occur in resource-limited settings. Obtaining the necessary equipments, qualified personnel, maintaining proper ventilation systems, and biomedical waste management can be difficult. Maintaining an uninterrupted flow of laboratory reagents, consumables, and BSL-3 PPEs was crucial for the operations. However, remote locations often faced delays and shortages due to logistical constraints, requiring thorough planning and contingency measures. Finally, developing robust emergency response plans and conducting regular drills were essential for ensuring the safety of laboratory personnel and the surrounding environment. Being prepared for potential incidents such as equipment failures or accidental exposures required diligent planning and preparation. Despite these challenges, our efforts to establish a laboratory for aliquoting nipah suspected samples in a remote setting were driven by a commitment to public health and safety.

While a BSL-2 lab can be used for diagnostic purposes, the ideal scenario would be a MBSL-3 laboratory for maximum safety. The development of the MBSL-3 laboratory following the COVID-19 pandemic in 2022 was a significant milestone for India, as it included all of the containment elements on wheels.

#### Zones and overview of the MBSL-3 Laboratory of ICMR

##### 2.2.1. Zones

The work area of MBSL-3 laboratory is divided into four zones consisting of Zone-1 [driver and outer change room]; Zone-2 [shower and inner change room], Zone-3 [main laboratory], Zone-4 [material staging and decontamination area]. The zone-wise major installations in MBSL-3 laboratory has been described in Table 1 and Figure 1.

**Figure 1:**
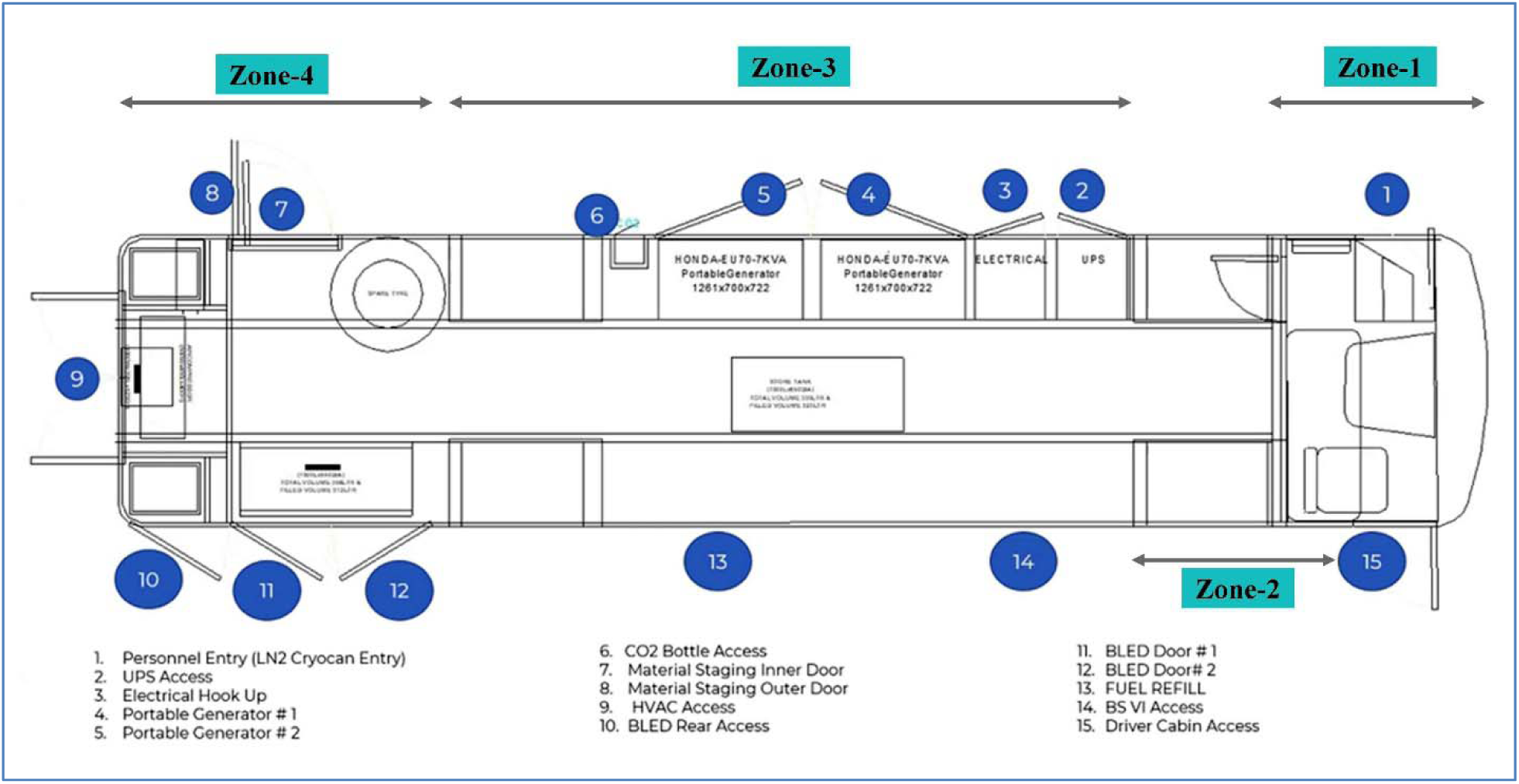
Zone-wise layout of the MBSL-3 laboratory.

**Table 1.** The zone-wise major installations of MBSL-3 laboratory.

| **Zones** | **Area in MBSL-3 laboratory** | **Major installations** |
| --- | --- | --- |
| Zone-1 | Driver area and Outer Change room | • Vehicle performance software and geo-location<br>• Door Gaskets |
| Zone-2 | Shower area and Inner Change Room | • Shower system<br>• Door interlocking system<br>• Emergency switches<br>• Inbuilt heater |
| Zone-3 | Main Laboratory | • Power Supply<br>• HVAC<br>• Biological safety cabinets [Class II A2]<br>• Intelligent programmable logic controller system |
| Zone-4 | Material Staging and Decontamination area | • Pass Box [400*400*400 mm]<br>• Autoclave [140 litres]<br>• BLED [225 litres]<br>• Air Compressors<br>• Supply water tank [420 litres]<br>• Drain Tank [425 litres] |

##### 2.2.2. Overview of the MBSL-3 laboratory

###### 2.2.2.1. Power supply

The power supply of the MBSL-3 laboratory is mainly through the direct electric supply [125 Amp] or Diesel Generator [62.5KVA]. The consumption of the power by MBSL-3 laboratory is monitored and recorded using energy consumption meter. In case of the sudden power failure while working in MBSL-3 laboratory, the back-up of the laboratory equipment is provided through uninterrupted power supply (UPS) system [10 KVA] and two petrol generators (PG) 1 and 2 of 19 litres capacity [7.5 KVA, EU70 Honda, Victoria, Australia].

###### 2.2.2.2. HVAC

The HVAC system maintains the unidirectional negative pressure in the MBSL-3 laboratory from entry side to laboratory area with temperature ranging from 18°C to 27°C. The cascade of pressure increases from outer change room to laboratory area with steps of 0 Pascal (Pa), −10 Pa, −25 Pa, −40 to −60 Pa. The air supply is 100% filtered with the 30 air change per hour. The air supply is purified with three filters including pre-filter [EU-6], intermediate filter [EU-8] and single HEPA filter [EU-14]. Similarly, the exhaust air is passed through HEPA filter (100%).

###### 2.2.2.3. Pass Box

A dynamic pass box (400×400×400 mm) is located at the material staging area with ultraviolet rays (UV) cycle. The pass box can be utilized for entry of sample boxes and laboratory supplies.

###### 2.2.2.4. Rapid Transport Port (RTP)

The RTP [70 cm diameter] with attached biohazard bag with ‘O’ ring [45 cm diameter] is provided at the emergency exit. Post inactivation of the samples, this port is utilized for directing the sample outside the MBSL-3 laboratory for further processing.

###### 2.2.2.5. Biological Safety Cabinets (BSC)

Two BSCs of Class II A2 [3 feet, BioKlenzaid, India] has been provided in the laboratory, one for handling the clinical specimens and other for the clean work.

###### 2.2.2.6. Entry-Exit shower system

The entry to MBSL-3 laboratory is biometric controlled and through the outer change room. The outer change room is provided with storage arrangements for personnel clothes and for donning of the PPEs. The personnel exiting the laboratory post completion of work is desired to perform surface disinfection on PPEs and then enter the inner change room. The inner change room is provided with the storage compartment with charging ports and UV disinfection system for the Powered Air Purifying respirator (PAPR) unit [3M, India] with HEPA filter. The liner port [70.10 cm diameter] with attached biohazard bag with ‘O’ ring [50 cm diameter] is provided in the inner change room for the discarding the PPEs (coverall, gloves, head cap and shoe-cover) which can later be collected for autoclaving. The personnel have to complete the shower cycle before exiting the laboratory. The drain of the shower is connected to drain tank which is attached to the BLED.

###### 2.2.2.7. Water supply and drain tank

There is a provision of supply water tank (420 litres) with inbuilt heater. The drains from the wash basin of the laboratory and from the shower are connected to the drain tank (425 litres). Once the drain tank is full, the water automatically gets transferred (sensor operated) to BLED for sterilization cycle.

###### 2.2.2.8. Autoclave

A double door autoclave (140 litres) is provided at the material staging area for sterilization of solid wastes generated in the MBSL-3 laboratory. The steam cycle along with the vacuum leak test [before each run] is utilized for sterilization and validation is done using biological [Sterkind, MicroBiotech Inc, India] and chemical indicators [3M, India].

###### 2.2.2.9. BLED

The single BLED tank (225 litres) is provided below the material staging area (Zone 4). There is provision of biological indicator [Sterkind, MicroBiotech Inc, India] port on the dish-end plate for the validation of the sterilization cycle. After the completion of sterilization cycle, the effluent is discharged into sewage treatment plant post attainment of 37°C temperature.

The major installations like autoclave, pass box, BLED, and vehicle performance system, shower are operated through programmable logic controller (PLC) [Siemens SIMATIC Step 7 1200 software, Germany] and intelligent monitoring [IM] panel [Siemens ET200SP, Germany].

###### 2.2.2.10. Decontamination of laboratory using Hydrogen peroxide (H2O2) fogger

The MBSL-3 laboratory is provided with H_2_O_2_ fogger for the fumigation and laboratory decontamination. A 2.5 litres of H_2_O_2_ [30% ready to use solution; Thermo Fisher Scientific India Pvt Ltd, India] is used in single cycle of fumigation for 20 min with the holding time of 1 hour and validation is done using chemical indicator [Rep CI115 Steritech, USA].

###### 2.2.2.11. Air Compressors

The totals of five air compressor units [BioKlenzaid, India] are provided in MBSL-3 laboratory.

###### 2.2.2.12. Cold storage systems

The laboratory has facility for the storage of clinical specimens and diagnostic reagents including deep freezers [[-20°C (98L) and −80°C (200L), Thermo Fisher Scientific, USA], refrigerator [+4°C (50L, Elanpro, India] and ice flaking machine. Additionally, it has backup of liquid nitrogen container [-196°C (47L), INOXCVA IX 47, India].

###### 2.2.2.13. Vehicle performance software with intelligent control automation and geo-location

The MBSL-3 laboratory is provided with logic and instruction controllers required for data acquisition and monitoring through IM automation system of power panel [Siemens ET200SP, Germany] with PLC [Siemens SIMATIC Step 7 1200 software, Germany].

The data on the critical parameters of the major installation can be monitored and recorded through human machine interface (HMI) system [Siemens HMI TP700, Germany] inside the laboratory and vehicle performance software [ZENON SCADA software, Austria].

The global positioning system (GPS) [Intangles Lab Pvt Ltd, India] has been installed in the Zone-1 of the MBSL-3 laboratory which enables the live tracking and the movement of MBSL-3 laboratory.

## 3. Results

### 3.1. Rapid deployment of MBSL-3 laboratory during nipah virus outbreak 2023

#### 3.1.1. Deployment and operationalization of MBSL-3 laboratory

As per the directives from ICMR headquarters, a team of trained scientists and technologists from ICMR-NIV, Pune along with engineer and driver from Klenzaids Contamination Control, Pvt Ltd, were deputed with MBSL-3 laboratory for initiating the onsite diagnosis of nipah virus at Kozhikode, Kerala on September 13, 2023 at 2:00 hrs Indian standard time (IST) (Figure 2). The MBSL-3 laboratory was transported to government medical college (GMC), Kozhikode, Kerala through the road-ways covering a distance of 1050 kms and it reached the destination on September 14, 2023 at 10:00 hrs IST. The travel duration was approximately 32 hours considering the maximum speed limit of 70 km/hour due to the weight of MBSL-3 laboratory [17 tons] and resting phase of the driver. With the support from GMC, Kozhikode, and state government of Kerala, it took additional ten hours to fully operationalize the MBSL-3 laboratory on the main electric power supply [125 Amp] from the electric sub-station. Additionally, the laboratory was supplied with water from water pumping station and the arrangements were made for discharging the treated effluent from BLED tank. In case of sudden power failure, the MBSL-3 laboratory was well-equipped with UPS and PG that could power the critical major installations and equipment for one hour. The onsite field nipah virus diagnosis was initiated after the complete operationalization and stabilization of the laboratory equipment on September 14, 2023.

**Figure 2.**
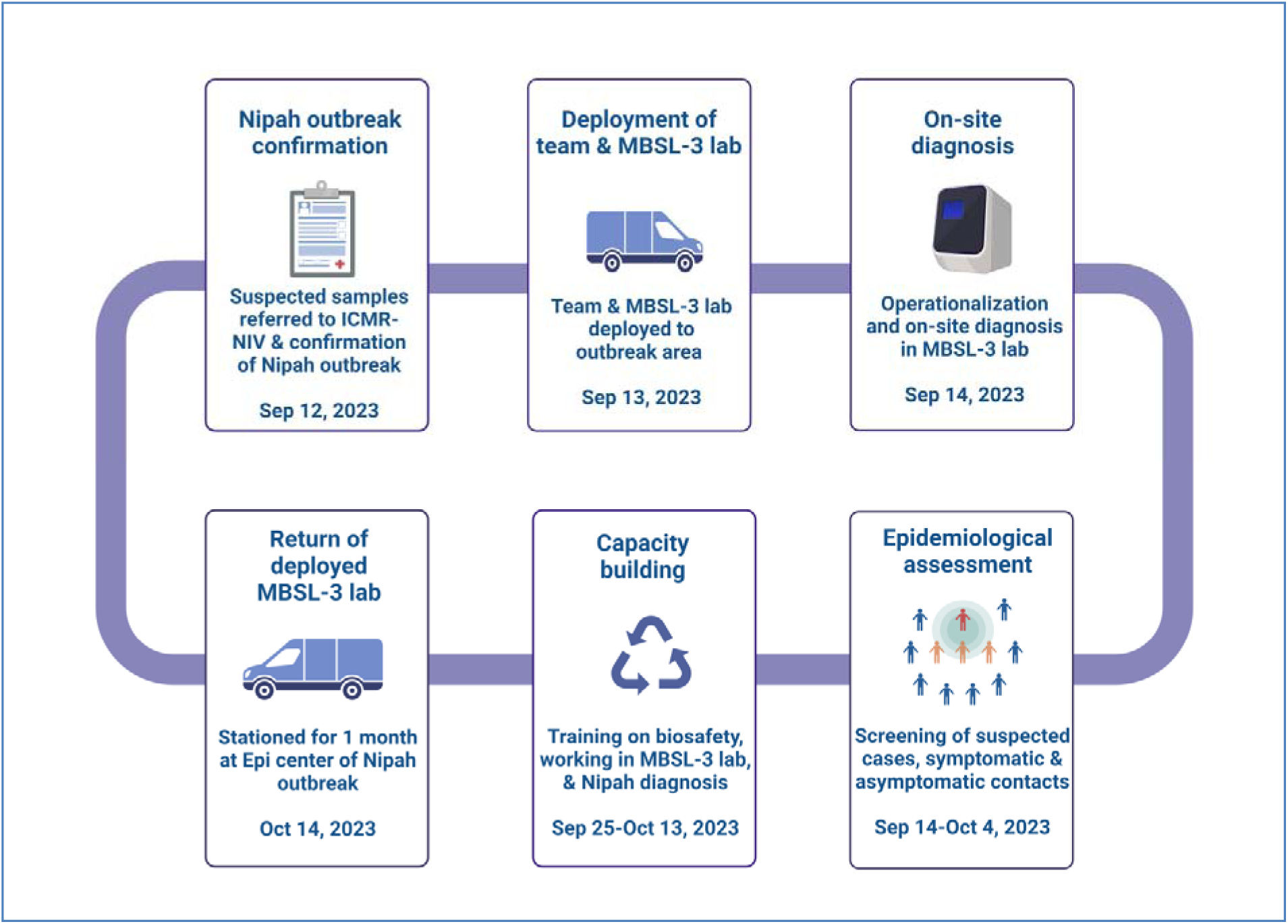
Series of events during the nipah virus outbreak response in Kozikode, Kerala.

#### 3.1.2. Processing of clinical specimens, RNA extraction and PCR

##### 3.1.2.1. Laboratory supplies and consumables

All the laboratory supplies and consumables required for processing of the clinical specimens, diagnosis, and decontamination of biomedical waste alongwith the biological and chemical indicators were transported from ICMR-NIV, Pune to GMC, Kozhikode on September 13, 2023. After the complete operationalization and stabilization of the MBSL-3 laboratory and thorough surface decontamination, the laboratory supplies and consumables were shifted and placed at the desired locations to initiate the testing.

##### 3.1.2.2. Clinical specimens receipt and processing

The specimens were handled and processed following the strict biosafety protocols, BSL-3 practices, and the SOPs. The PPEs used were coverall with booties, pair of dedicated laboratory boots, shoe cover, PAPR with hood, head cap and double pair of gloves. The standard triple packaging method was used for delivering the clinical specimens from the isolation ward to the MBSL-3 laboratory. The surface of the specimen boxes were disinfected using 1% benzalkonium chloride [3M, India]. The dynamic pass box was used for the entry of the box containing the specimens inside the laboratory. The specimens were handled in the dedicated class II A2 BSC and the individual specimens were aliquoted in different volumes including an aliquot for RNA extraction in inactivating agent (Figure 3). The other aliquots were stored in −80°C deep freezer of MBSL-3 laboratory and later were transported to ICMR-NIV, Pune in dry ice post surface disinfection.

**Figure 3.**
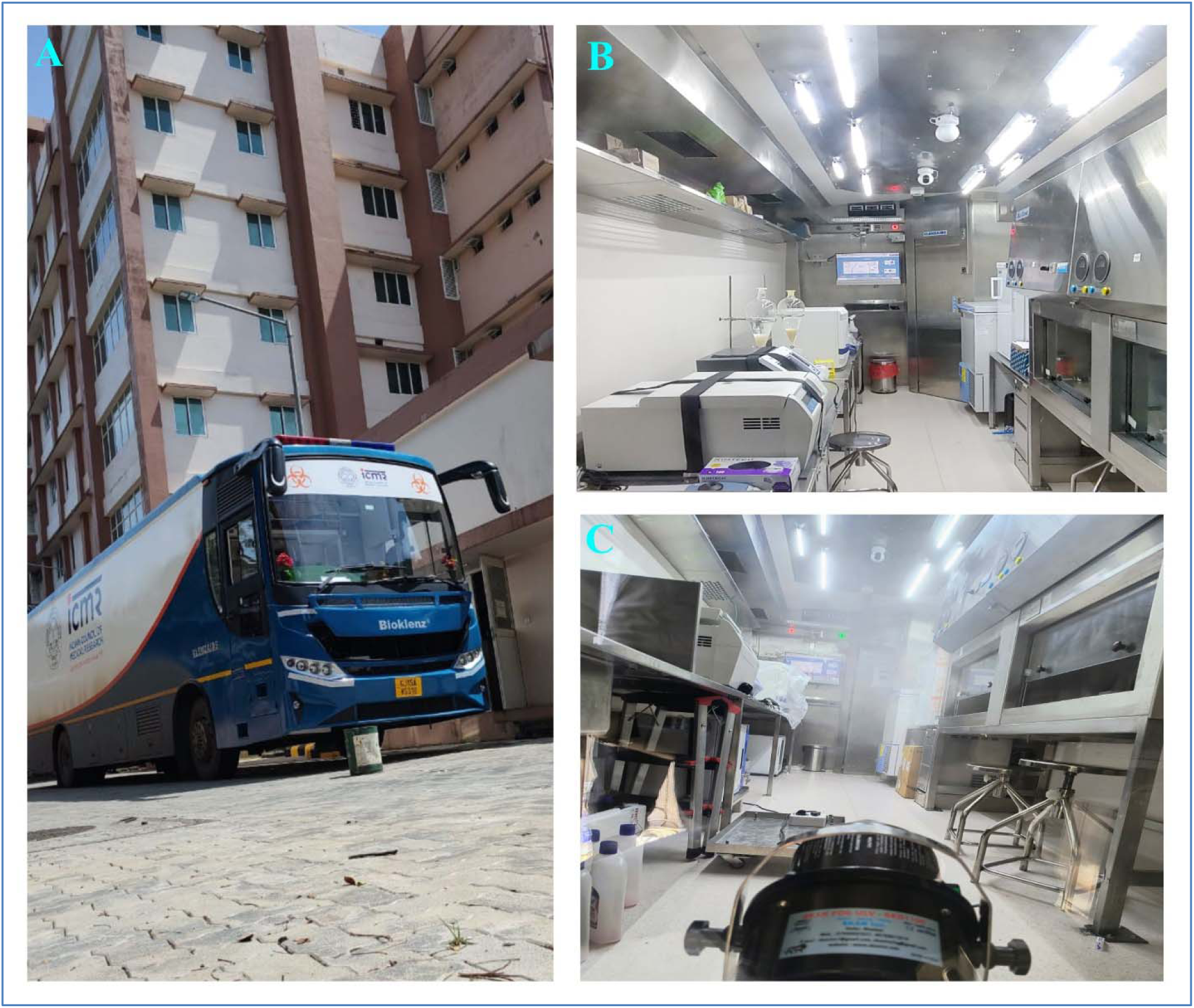
Representative images of the onsite activities of MBSL-3 laboratory during nipah outbreak 2023: A) MBSL-3 laboratory stationed in the campus of Government Medical College, Kozhikode, Kerala, India. B) The inside view of the MBSL-3 laboratory. C) Decontamination of the MBSL-3 laboratory using H2O2 fogger.

##### 3.1.2.3. RNA extraction and Real time RT-PCR

KingFisher™ Duo Prime Purification System (Thermo Fisher Scientific, USA) has been installed in MBSL-3 laboratory which is based on magnetic-bead based nucleic acid extraction, ensuring reproducible recovery of high-quality nucleic acid for a range of downstream applications, such as sequencing and qPCR. Commercially available HiPurA® Viral RNA Purification Kit [HiMedia, India] was used for nucleic acid extraction from the clinical specimens as per the manufactures instructions. The collected specimen were handled in BSC and inactivated using the 560 µl of lysis buffer containing Proteinase K, carrier RNA and magnetic beads. Plate containing the samples, wash buffer, 70% ethanol and elution buffer was placed in the KingFisher™ Duo Prime machine as per the manufacturer’s instructions. Extracted RNA was immediately used for nipah virus rRT-PCR. Evaluation of the efficiency of the extraction system was performed before the deployment of MBSL-3 laboratory using known positive and negative specimens in duplicate and the extracted nucleic acid were tested and validated by nipah specific rRT-PCR.^8^

The rRT-PCR assay was performed on extracted nucleic acid using the TaqMan based assay targeting nipah NP gene with RNase P as an internal target control.^8^ RNA was amplified using the TaqMan™ Fast Virus 1-Step Master Mix [Thermo Fisher Scientific, USA] and 40-cycle assays were run using Applied Biosystems 7500/7500 Fast rRT-PCR System [Applied Biosystems, USA]. The specimens were considered positive if cyclic threshold (Ct) value of nipah N gene was ≤ 38. In contrast, the specimens were considered negative if there was no amplification with the Ct value undetermined. In both the scenarios internal control Ct value should be ≤ 35 indicating proper extraction of nucleic acid from clinical specimens. The turnaround time for the nipah diagnosis on referred clinical specimens was around 4 hours.

The clinical specimens of a total of 165 suspected cases and contacts were screened by rRT-PCR at the MBSL-3 laboratory during September 14 to October 4, 2023. These included the symptomatic contacts (n=35; high risk-30, low risk-05); and the non-epidemiologically linked suspected Nipah cases (n=130).^9^ In the recent nipah virus outbreak, a total of six cases were confirmed, of which initial five cases were tested at apex laboratory of ICMR-NIV, Pune and one positive case was tested at MBSL-3 laboratory.

##### 3.1.2.4. Reporting

All the suspected cases and the clinical specimens were provided with the unique identifiable number from the field set up. The nipah virus suspected case investigation form was filled by the doctors and were accompanied with the clinical specimens. All the nipah diagnostic test results were communicated to the authorities and officials from hospital, state health ministry, and state integrated disease surveillance program, National Center for Disease Control, Emergency Medical Relief department, and ICMR. The report included the field and hospital identification, name of referral hospital, name of the suspected case, gender, age, and results of nipah rRT-PCR assay.

##### 3.1.2.5. Communication and surveillance system inside the MBSL-3 laboratory

The walkie talkie [Vertel Digital Pvt Ltd, India] has been provided for the communication with laboratory personnel working in the MBSL-3 laboratory from outside the laboratory. The MBSL-3 laboratory is well equipped with the closed circuit television camera (CCTVs) for the real time surveillance of the laboratory activities. Similarly, the intelligent control automation system was provided with the local area network (LAN) ports for sharing the data of the major installations and equipment. The test results and reports were communicated through the laptop placed inside the MBSL-3 laboratory.

### 3.2. Decontamination and biosafety and biorisk management in MBSL-3 laboratory

#### 3.2.1. Movement of laboratory personnel

No more than three personnel were working inside the laboratory at a particular time to avoid the cluttering and disturbance in the air exchanges in the laboratory. The duration of work was restricted to a maximum of six hours for one team which would later be replaced by a second team of three personnel. The personnel entry was done from the outer change room through biometric access following the defined and validated entry-exit SOPs. Before entering the laboratory, the personnel had donned the laboratory scrubs and PPEs.

After completion of the work, the PPEs were surface disinfected using 1% benzalkonium chloride and after contact of 10 min, the person exited the laboratory into the inner change room. The PPEs were then doffed as per the SOP and discarded in the dedicated PPE discard liner port. The PAPR units were surface disinfected and kept for charging for further usage in the defined cabinets with the facility of UV sterilization. The personnel then used to enter the shower room and after completing the three cycles of shower entered the outer change room and then exit the laboratory.

#### 3.2.2. Solid-waste management

All the solid bio-waste was surface disinfected and segregated at the point of generation. The double walled autoclave steam sterilization cycle was run on daily basis at night to decontaminate the solid waste. The waste was later collected in the morning hours before the start of the testing procedures and later discarded as per the state biomedical waste management guidelines. The chemical indicators and the biological indicators (*Geobacillus stearothermophilus*) were used for the sterilization cycle validation for each run.

#### 3.2.3. Liquid-waste management

The water waste generated inside the MBSL-3 laboratory was collected in the drain tank which automatically shifts into the BLED tank as per the commands generated from the sensors. When the sensor indicator of the BLED tank showed the ‘high’ load, the sterilization cycle was initiated. Considering the work load, the BLED sterilization cycle was also done on a daily basis after the completion of the autoclave cycle at night hours. The biological indicator ampoule (*Geobacillus stearothermophilus*) was placed in the port provided on dish-end plate for the validation process of each run. The treated effluent was later discharged in the sewage treatment plant in the morning hours once the water temperature reached 37°C.

#### 3.2.4. Air flow

The laboratory is designed in such a way that there is always an unidirectional air flow with a cascade of negative pressures from outside to inside. All the air entering and exiting the laboratory were HEPA filtered. The air supply was controlled with 30 air changes per hour.

#### 3.2.5. Surface disinfection and fumigation

All the stainless-steel surfaces inside MBSL-3 laboratory were disinfected using 1% benzalkonium chloride solution, followed by dry mopping after a contact time of 20 minutes. Later all the surfaces were disinfected with 70% ethyl alcohol swabs and allowed to air dry. The MBSL-3 laboratory was fumigated using 30% hydrogen peroxide (H2O2) solution (ready to use) by fogger after every 72 hours (Figure 3). The chemical indicators were used to validate the effective decontamination after vaporized H2O2 fumigation cycle.

#### 3.2.6. Health monitoring of the laboratory personnel

Each deployed staff was monitored for health complaints on a daily basis and was given sufficient rest and personal space in between shifts. None of the deployed personnel had reported any medical illness during and post deployment.

### 3.3. Checklist for the MBSL-3 deployment

The checklist helps in the identification of various response, considerations and arrangements required for timely coordination for the movement of MBSL-3 laboratory and required laboratory investigation on site (Table 2). The primary utilization of RRML would focus on field logistics including custom clearance, transport of materials and reagents, laboratory information, and testing quality management, with enhanced biosafety and biosecurity.

**Table 2.** Key parameters and checklist for deployment of MBSL-3 laboratory.

| Parameters | Pre-Deployment [Pre-Mission] | Deployment [Mission] | Post-Deployment [Post-Mission] |
| --- | --- | --- | --- |
| **Health of the personnel** | • Deployment of only trained personnel<br>• Fitness of the Deployed staff including mental health assessment<br>• First aid box with Automated External Defibrillator (AED) status | • Health Monitoring throughout deployment<br>• Shift duties and rest period<br>• Information about the nearest health care facility<br>• Basic Life Support trained staff for medical emergency management | • Quarantine policy as per the government norms<br>• Health monitoring<br>• First aid box with Automated External Defibrillator (AED) status |
| **Route of mobilization and stationing of MBSL-3 laboratory** | • Odometer reading<br>• Tyre pressure status<br>• Diesel Exhaust Fluid Level (AdBlue)<br>• Engine function<br>• Diesel level<br>• Geo location monitor<br>• Assessment of road and highways for safe movement<br>• Energy Meter reading<br>• Road Permit/Air permit<br>• Vehicle light and indicator status<br>• Wind Shield cleaner water level | • Energy meter reading<br>• Availability of water supply, drain discharge facilities, electric supply (direct or DG generator) and shades to cover MBSL-3 laboratory<br>• Diesel level<br>• Tyre pressure status<br>• Vehicle light and indicator status | • Odometer reading<br>• Tyre pressure status<br>• Engine function<br>• Diesel Exhaust Fluid Level (AdBlue)<br>• Geo location records<br>• Energy Meter reading<br>• Diesel level<br>• Vehicle light and indicator status<br>• Wind Shield cleaner water level |
| **Major Installations functioning** | • PLC system working<br>• HVAC<br>• BLED Autoclave<br>• Pass box<br>• Entry-Exit Shower<br>• H2O2 fogger<br>• Bio-metric access control | • Utilization of all the major installations and record generations | • Recording the data of the major installations<br>• Servicing and calibration of the major installations, if needed |
| **Approvals** | • From competent authority<br>• MBSL-3 laboratory movement approvals<br>• Road Traffic Office pass/Air movement pass<br>• Vehicle documents | • From competent authority for stationing the MBSL-3 laboratory<br>• Information to local police<br>• Information to local hospital | • Record keeping the approvals<br>• Records of all the bills |
| **Equipment check** | • Fire extinguishers refilled and status | • Fire extinguishers trained staff | • Fire extinguishers refilled and status |
|  | • Electric connections removed<br>• PG emptied<br>• Walkie talkie<br>• CCTV | • Electric connections done<br>• PG refilled<br>• All equipment functional [rest phase post movement (4 hours)]<br>• Walkie talkie<br>• CCTV | • Electric connections done<br>• PG refilled<br>• Check for the equipment functionalization<br>• Walkie-talkie servicing<br>• CCTV recordings and photos backup |
| **Supplies** | • H<sub>2</sub>O<sub>2</sub> liquid<br>• Chemical indicator<br>• Biological Indicator<br>• Reagents for testing<br>• Laboratory supplies<br>• PPEs including PAPR<br>• Scrub, socks, towels<br>• Disinfectant, decontamination solutions<br>• Water pipes<br>• ETP drain assembly<br>• Critical extra spares<br>• Loading the MBSL-3 laboratory tools<br>• Power cables | • Keep the material at secured places and utilize it as needed in field<br>• Waste management<br>• Decontamination procedures | • Unloading of laboratory supplies<br>• Cleaning of MBSL-3 laboratory<br>• Servicing of major installations<br>• Unloading of the critical spares and tools<br>• Check the inventory and records |
| **Laboratory testing Quality Assurance and Quality Control(QAQC)** | • All the desired laboratory diagnostic tests to be performed to check the quality of reagents and working of equipment<br>• Calibration and maintenance of equipment<br>• Proficiency test records for competency of the laboratory personnel | • Continuous monitoring of the test and adherence to the SOPs<br>• Reporting of non-conformities if any | • QAQC |
| **Laboratory Data Quality Management** | • Standard Operating procedures (SOPs) for diagnostic test and equipment<br>• Data protection and archiving<br>• Check the training records of personnel<br>• Log books | • Adherence to SOPs<br>• Coding of specimens<br>• Patient information<br>• Inventory of specimens and reagents<br>• Data and test reporting<br>• Cloud storage<br>• Daily log book entries | • Data protection and archiving<br>• Inventory check<br>• Storage of specimens in the repository of ICMR-NIV, Pune |
| **Journey status** | • Planning and execution and departure date and time | • Duration of stay with dates and time | • Planning and execution and arrival date and time |

Intermission period to be utilized for the shutdown and maintenance of MBSL-3 laboratory, training of the personnel with mock drills and simulation exercises as well as daily monitoring of the engineering parameters. A preventive annual medical examination of the personnel including in-depth psychological evaluation, including vaccination are important and crucial considerations for the deployment in outbreak setup.

## 4. Discussion

Government, medical, and public worked in tandem and were quick to come up with an effective response for the Kerala outbreak. This response included the deployment of the MBSL-3 laboratory which was crucial. It significantly helped public health authorities to make quick decisions by reducing the amount of time required to process samples. Officials went ahead to deploy the laboratory directly to the affected area to conduct on-site diagnosis to enhance the turn out time for reporting. The availability of MBSL-3 laboratory in India not only enhances the outbreak response but also strengthens the public health delivery capabilities of the country. In determining the pathogen risk groups, a risk assessment should be undertaken that takes into account the deployment scenarios for outbreak responses. Workforce capacity building to work in RRML is crucial for enhancing the teams required for deployment in both non-emergency and emergency situations. Staff allocated to an RRML should be appropriately trained in every pertinent field related to MBSL-3 laboratory.^1,2,10^

World Health Organization (WHO) has also taken up the initiative for the scenario-based table-top exercises for the deployment of RRML with the opportunity to explore the avenues for enhancing collaborations and coordination.^1^ This RRML can be utilized in the outbreaks/epidemics and pandemics of zoonotic potential, one health, and potential biological threats.^11^ This MBSL-3 laboratory can also be utilized for the outbreak responses in the South East Asian Countries. Logistics for the overseas deployment of MBSL laboratory demands meticulous preparation for providing immediate delivery of diagnostics and therapeutic countermeasures as part of the outbreak response. MBSL provide a vital resource at places far from specialized laboratories, as well as are easier to locate and relocate as needed during an outbreak.^11^ The major limitation of deploying MBSL-3 laboratory is the breakdown that could happen at any time point during field outbreak investigations which affect the operationalization and refute the purpose of deployment. Deploying a trained engineer with the testing team would be desirable as done during the nipah outbreak 2023. Availability of the contingency amount at all the time during deployment would help in daily small wear and tear maintenance. Apart from this, there is always a perceived risk regarding the civil unrest and threats in the dangerous outbreak areas which pose security threat to the deployed personnel, MBSL-3 laboratory and the biological specimens.

## 5. Conclusion

The nipah outbreak in Kozhikode, Kerala 2023, posed significant challenges to public health. The MBSL-3 laboratory was deployed as an integral part of this outbreak containment responses. The utilization of this MBSL-3 laboratory during nipah outbreak in Kerala, India is an exemplary example of a rapidly deployable, and sophisticated infrastructure adding strategic advantages to meet the epidemiological challenges. It’s a new paradigm to the India’ fore front battles against the high risk pathogen outbreaks with quick, effective and timely response.

## Ethical approval

The study was approved by the Institutional Human Ethics Committee of ICMR-NIV, Pune, India under the project ‘Sustainable laboratory network for monitoring of Viral Haemorrhagic Fever viruses in India and enhancing bio-risk mitigation for High risk group pathogens’ [NIV/IEC/March/2021/D-9 dated April 9, 2021]. No patient related data has been included in this manuscript.

## Author Contributions

PDY, RRS, DYP, AMS, SM, and NG supervised and co-ordinated the nipah outbreak response from ICMR. RRS, and DYP established the field MBSL-3 laboratory and provided the onsite diagnosis. PDY, AMS, DYP, RRS, and SM supervised the laboratory investigations and quality assurance and quality control. NG provided administrative support for deployment of MBSL-3 laboratory. DTM, PDY, RRS, DYP, and AMS contributed in validation of MBSL-3 laboratory. PDY, RRS, DYP, and AMS contributed to data collection, interpretation, writing and critical review. All the authors have contributed equally in reviewing and editing the final manuscript.

## Data Availability

All the data related to this study has been included in the manuscript

## Acknowledgement

Authors take this opportunity to convey thanks and appreciation to all individuals who have contributed immensely towards the Nipah virus outbreak response and containment measures in Kozhikode district, Kerala state, India during September 2023 including Smt. Veena George, Hon’ble Minister for Health and Family Welfare, Kerala, and Shri. Mohammed Hanish, Principal Secretary for Health, Kerala. The authors are thankful to Dr. Rajiv Bahl, Secretary Department of Health Research and Director General, Indian Council of Medical Research (ICMR), New Delhi for his constant support, guidance and motivation. Authors would like to thank Dr. Sheela Godbole, Director In-charge, ICMR-National Institute of Virology (NIV), Pune for administrative approvals required for the outbreak responses. The authors would like to thank Dr. Thomas Mathew, Director of Medical Education; Smt. A Geetha, District Collector Kozhikode; Dr. Shaji Cheriya Koroth, District Program Manager and Dr. Neelakandhan Asokan, Principal, Government Medical College (GMC), Kozhikode, for providing the logistic supports for the onsite stationing and smooth functioning of MBSL-3 laboratory at GMC campus. Authors extend sincere appreciation for the diligent work put in by all members of the scientific and technical staff deputed for on site for laboratory testing at MBSL-3 laboratory from ICMR-NIV, Pune and ICMR-NIV, Kerala unit including Dr. Anukumar Balakrishan, Dr. Kannan Sabarinath PS, Dr. Ullas PT, Dr. Siba S, Dr. Surendra Kumar, Mr. Ratnadeep More, Mr. Rameshwar Khedekar, Mr. Deepak Mali, Mr. Jijo Koshy, Mr. Dinesh Kumar Singh, Mr. Sunil Shelkande, Mr. Hitesh Dighe, and Mrs. Najiya KV. Authors extend sincere gratitude for the engineering support by Mr. Rajeshwar Dhodi and driver of MBSL-3 laboratory, Mr. Kundan Gupta from Klenzaids Contamination Control Pvt Ltd, Mumbai for onsite support during MBSL-3 laboratory deployment. We also acknowledge the excellent technical support from Dr. Rajlaxmi Jain, Mr. Ganesh Chopade, Mr. Sanjay Thorat, and Mr. Madhav Acharya for transportation of reagents, PPEs and other logistics at the site of outbreak. We extend the sincere thanks to the support for the laboratory diagnosis and sample logistic support from the scientific and technical team at Regional Virus Research and Diagnostic laboratory at GMC Kozhikode lead by Dr. Anitha PM, Professor and Head Department of Microbiology. Authors are thankful to Dr. Shailesh Pawar, Scientist-F and Officer In-Charge, ICMR-NIV Mumbai Unit; Dr. Jayati Mullick, Scientist-F and group leader, Polio virus group; Er. Ajay Khare, Senior Technical Officer and their teams [Mr. Dinesh Singh, Mr. Sachin Keng, Mr. JPN Babu, Mr. Gajanan Ghogare, Mr. Shashikant Surbhaiya] and team from Maximum Containment Facility [Er. Mayur Mohite, Er. Nand Kumar, Er. Vishal Gaikwad, Mrs. Triparna Majumdar, Mr. Deepak Mali] of ICMR-NIV, Pune for the validation of indigenous MBSL-3 laboratory. Authors extend the scientific support for validation of MBSL-3 laboratory from Dr. Jitendra Narayan, ICMR New Delhi. Authors extend sincere thanks to team from Klenzaids Contamination Control Pvt Ltd, Mumbai (Mr. Hamish Sahani, CEO; Mr. Sanil Diwakaran, Mr. Biju Matthews, Mr. Jacob Vasantraj, Mr. Tushar Banerjee, Mr. Yogendra Gondane) for the development of MBSL-3 laboratory and supporting ICMR-NIV, Pune team for the validation work.

